# Exploring provider preferences in the design of HIV treatment packages integrating long-acting injectable antiretroviral therapy in New York Ryan White Part A medical case management programs: A discrete choice experiment

**DOI:** 10.64898/2026.04.22.26351494

**Authors:** Rebecca Zimba, Elizabeth A. Kelvin, Sarah Kulkarni, Jennifer Carmona, Tigran Avoundjian, Connor Emmert, Meghan Peterson, Mary Irvine, Denis Nash

**Affiliations:** CUNY Institute for Implementation Science in Population Health, City University of New York, New York, NY, USA; Department of Epidemiology and Biostatistics, CUNY Graduate School of Public Health and Health Policy, City University of New York, New York, NY, USA; Donald and Barbara Zucker School of Medicine at Hofstra University/Northwell Health, Hempstead, NY, USA; Division of Disease Control, New York City Department of Health and Mental Hygiene, New York, NY, USA; Bureau of HIV, Hepatitis, and Sexually Transmitted Infections, New York City Department of Health and Mental Hygiene, New York, NY, USA

**Author notes:** **Corresponding Author:** Rebecca Zimba. **Authors’ note:** As of the time of this publication, Tigran Avoundjian was affiliated with Public Health -- Seattle & King County, Washington, USA, and Jennifer Carmona was affiliated with the Bureau of Equitable Health Systems at New York City Department of Health and Mental Hygiene, New York, NY, USA.

**Keywords:** HIV, Antiretroviral therapy, Long-acting injectables, Healthcare providers, Discrete choice experiment, New York

## Abstract

**Introduction:** Understanding provider preferences for the design of HIV treatment packages could enhance the implementation of programs to support the adoption of long-acting injectable antiretroviral therapy (LAI ART) by people living with HIV who are interested in initiating this treatment modality.

**Methods:** We recruited providers from New York City (NYC), Rockland, Putman, and Westchester County Ryan White Part A Medical Case Management (MCM) programs to complete a discrete choice experiment (DCE) containing twelve tasks with two alternatives and an opt-out option, with additional survey questions about implementation readiness and choice motivations. The alternatives included four attributes—Type of ART Medication (monthly or bimonthly LAI ART), Service Location and Mode, Support for Clients, and Rewards for Clients— with 2–4 levels each. We ran latent class multinomial logit analyses (LCA) with 1–5 classes to estimate preferences and explore hypothesis-free preference heterogeneity. We estimated attribute influence using relative importances and preferences using zero-centered part-worth utilities for each level.

**Results:** One hundred seventy-seven providers completed the survey (July 2022–January 2023). About half (52%) were 40–59 years old, 72% identified as women, and the plurality (41%) identified as Latino/a. We chose the two-group LCA solution. Bimonthly LAI ART was preferred over monthly LAI ART overall and in both groups. Group 1 (n=45) preferred more traditional adherence supports (e.g., injections at the clinic by appointment, injection appointment reminders) whereas Group 2 (n=132) preferred more client-centered supports (e.g., injections at home by appointment, free transportation to injection appointments if at a clinic). Both groups preferred higher monetary value gift cards for clients for every on-time injection. The top-ranking motivations indicated that participants prioritized patient convenience over job satisfaction and administrative or financial feasibility for the agency. The scores for all implementation measures indicate readiness to implement LAI ART in both groups.

**Conclusions:** Our implementation science-focused study suggests that providers of MCM services in NYC and surrounding counties are motivated to offer services to support clients’ access and adherence to LAI ART. More work is needed to understand how programs have, in fact, integrated supports for LAI ART into their services.

## Introduction

Ryan White HIV/AIDS Program Part A (RWPA) medical case management (MCM) programs play a key role in addressing barriers to HIV care and antiretroviral therapy (ART) adherence for low-income, predominantly Black and Latino/a people with HIV (PWH).^1,2^ The New York City (NYC) Health Department manages RWPA MCM and “Care Coordination”^3-5^ programs in NYC and the suburban and rural Tri-County area north of NYC which provide services to >3,800 PWH each year. These programs have shown effectiveness in engaging PWH in care and treatment, but variability in clinical outcomes such as viral suppression (VS) and durable viral suppression (DVS) highlight the need for innovative strategies to increase long-term ART adherence.^6-10^

Merged surveillance data comparing NYC RWPA-enrolled PWH to other PWH in NYC who received care in 2022 showed that a higher proportion of RWPA clients were retained in care, defined as ≥1 viral load test, than other PWH in NYC (86% versus 78%).^11^ However, among those retained in care, RWPA clients had lower VS on last test than other PWH (83% versus 93%), and lower DVS on all tests (71% versus 86%).^11^ Given the high levels of HIV care and prescription coverage through NYS Medicaid^12^ and Uninsured Care Programs,^13^ most of the observed gaps in VS cannot be attributed to inadequate or differential financial resources available for ART access; in fact, higher proportions of Black (95%) and Latino/a (96%) than White (92%) PWH in NYC are on ART after HIV diagnosis.^14^ The drop-off appears related to ART use and adherence, as has been seen in other settings.^15,16^ These data underscore the potential to reduce VS disparities through a properly implemented regimen simplification combined with adaptations to current ART adherence support service strategies.

In January 2021, the US Food and Drug Administration (FDA) approved Cabenuva, which combines cabotegravir and rilpivirine in a long-acting injectable (LAI) formulation, as an alternative to daily oral ART regimens.^17^ In Phase III clinical trials, Cabenuva was found to be comparable in safety and efficacy and superior in patient satisfaction to daily oral ART.^18-21^ This treatment advance has the potential to resolve adherence barriers,^22^ close gaps in the HIV care continuum between care retention and VS, and reduce racial/ethnic disparities in VS, thereby accelerating the achievement of the United States Ending the HIV Epidemic (EHE) and UNAIDS 95-95-95 targets.^23-25^ However, optimizing the impact of this biomedical technology requires understanding not only preferences among PWH for specific ART regimen and treatment support options,^26-28^ but also provider views around treatment support packages that would be feasible and desirable within their service delivery contexts.^29,30^

One survey among Ryan White Part C program medical directors found that LAI ART was welcomed by 98% and considered appropriate by 86%, whereas 66% thought it would be easy to implement within their healthcare system and only 27% reported that their clinic was fairly to extremely ready to implement LAI ART.^31^ Other studies with clinical and non-clinical providers found that they saw LAI ART as shifting the responsibility for ART adherence from clients to providers, and they expressed concerns about missed injection appointments and billing/payment.^32-35^ Indicating the need to strengthen clinic systems to take on this new responsibility, and maximize its impact and equitable rollout, providers have suggested adding personnel, centralized tracking, storage capacity, expanded outreach, and mobile delivery of LAI ART.^32,33,35^

Implementation research is needed to ensure that LAI ART options are integrated in ways that meet both client and provider needs and preferences, which will likely vary by service setting and jurisdiction. Without provider buy-in and capacity for adding this ART delivery method as a choice within their service system, LAI ART will not be presented as a viable option to clients and uptake will remain low. We conducted surveys with discrete choice experiments (DCEs) among clients and providers in NYC’s RWPA MCM programs to elicit their preferences around integrating LAI ART as an option for clients. The surveys were part of a larger implementation-science study, Assessing Perceptions and Preferences around Long-acting Injectables (APPLI) in the Ryan White HIV/AIDS Program,^36^ focused on informing the delivery of LAI ART and related supports in safety-net service settings, for an equitable rollout of this major treatment advance. Here we report the results of the provider survey.

## Methods

### Survey design & pilot testing

Following best practices for DCE development,^37,38^ we convened focus groups to generate lists of possible elements to define the DCE—attributes, and categories within attributes called levels. From October–December 2021, we conducted two focus groups with MCM clients and three focus groups with MCM providers. During the development of the attributes and levels, we considered the focus group findings, feedback from the study’s Advisory Board (which we described in the study’s protocol paper^36^), and results from previous studies, including LAI ART clinical trials.^39,40^

The client-facing DCE assessed preferences for both daily pill ART and LAI ART.^28^ However, in the provider-facing DCE, we focused on eliciting preferences for supports and service options for LAI ART only, given that MCM programs and providers would be faced with integrating LAI ART (whereas clients could elect either regimen type). The provider-facing DCE contained four attributes: Type of ART Medication (monthly or bimonthly LAI ART), Service Location and Mode, Support for Clients, and Rewards for Clients. Each attribute contained two to four levels.

We used Sawtooth Software’s Lighthouse Studio to design and deploy the survey, which could be completed on any internet-connected device. We used the Balanced Overlap^41^ design option to create random combinations of levels for each alternative, with one prohibited pair of levels (“Injections by appointment at home” in the Service Location and Mode attribute was prohibited from appearing in an alternative with “Free transportation provided to get injections” in the Support for Clients attribute). Balanced Overlap obtains level balance and orthogonality without preventing the same level within an attribute from appearing randomly in both of the alternatives in choice tasks, which permits investigation of interaction between levels across attributes.^41^ The final provider DCE comprised twelve unlabeled choice sets with two alternatives and a None/opt-out option if neither alternative was acceptable (Supplemental Table 1 and Supplemental Figure 1).

Participants saw the following prompt: “Imagine that you had to choose between two treatment options with the features below for a MCM program client interested in long-acting injectable ART. Given your role in that program, select the option you would prefer the program to offer.” Based on feedback from pilot testing with ten clients in April 2022 and six colleagues at the Health Department in June and July 2022, we clarified question wording and provided additional instructions about scrolling to see all options when taking the survey on a mobile device. See Supplemental Table 2 for our completed DIRECT checklist.

In addition to the DCE, the survey contained questions allowing us to describe the study population: age group, gender identity, self-identified race and ethnicity, and duration of experience providing MCM services under RWPA. We also asked participants to separately rank the levels within each of the attributes, and we asked them to identify, from a list of pre-defined options as well as an “Other” option, the top three motivations for their ranking. We asked if they knew about LAI ART before the survey, and if so, both the sources of their knowledge and whether any of their clients had tried LAI ART. To assess the readiness of participants to incorporate supports for LAI ART into their programs, we included the implementation science Acceptability of Intervention Measure (AIM), Intervention Appropriateness Measure (IAM) and Feasibility of Intervention Measure (FIM) for LAI ART. Each measure is reported as the average of four items scored from 1 (“Completely disagree”) to 5 (“Completely agree”).^42^

### Study population & sample size

Eligible participants were affiliated with any of the 29 participating RWPA MCM programs in NYC (24 programs) or the Tri-County area (Rockland, Putnam, or Westchester, 5 programs) in job roles essential to MCM program delivery: prescribing clinicians; case managers/care coordinators; patient/peer navigators; health educators and coaches; and/or program administrators/managers. Hereafter we refer to all of these individuals as providers.

We used two methods to determine our sample size. First, we used Johnson and Orme’s formula “*n* ≥ (500*c*)/(*ta*)” to obtain the sample size needed to estimate main effects in a DCE with precision.^43,44^ The minimum sample size for the provider DCE was 83, given a maximum of four levels per attribute, 12 choice sets and 2 alternatives per set (excluding the none option). The second method we used set the sample size so that the estimated standard errors for the main effects in the DCE design were ≤0.05.^45^ Using Lighthouse Studio’s Test Design function,^46^ the largest standard errors were 0.09 with a sample size of 83. We adjusted our target sample size to 200 responses so that the estimated standard errors ranged from 0.03 to 0.05.

### Recruitment & data collection

Using existing NYC Health Department RWPA contract lists, the Health Department fieldwork team reached out to each MCM program to request a current list of names, titles/roles, and e-mail addresses of eligible providers. We sent the survey URL and unique login codes to the 341 eligible providers across all 29 programs. Before accessing the survey questions, providers were required to review and complete an electronic consent form. After submitting the survey, participants received a $25 gift card. The study methods were reviewed and approved by the Institutional Review Board of the New York City Department of Health and Mental Hygiene (protocol 20-096).

### Statistical analysis

Latent class multinomial logit analysis (LCA) was used to estimate main effects and to explore choice behavior heterogeneity within the survey sample without *a priori* hypotheses.^47^ We ran 1–5 class models, choosing the LCA solution based on change in log-likelihood, percent certainty (similar to McFadden’s pseudo-R^2^), Akaike’s Information Criterion (AIC), the size of each group, and the extent to which the differences between groups yielded practical interpretations.^48^ We estimated attribute influence using relative importance: the range of utilities of an attribute divided by the sum of ranges of utilities for all attributes, on the percent scale.^43,48^ We estimated preferences using zero-centered effects coded part-worth utilities for each level.^43,45^ We conducted sensitivity analyses excluding poor quality responses as gauged by response speed (faster response times correlating to poorer quality) and straight-lining (always selecting the left-hand option, the right-hand option, or the None/opt-out option),^49,50^ We described study participants overall and by LCA group using counts and percentages for categorical variables, and means with 95% confidence intervals (CIs) or medians with interquartile ranges (IQRs) for continuous variables. We conducted statistical testing of differences by LCA group; for categorical variables we used the Chi-square test or Fisher’s exact test, and for continuous variables we used the Wilcoxon rank-sum test. We used Lighthouse Studio’s Analysis Manager to generate the LCA solutions and sensitivity analyses, and R Studio 2025.05.0 build 496 (running R version 4.4.2) for all other analyses.

## Results

### Participant characteristics

From July 2022–January 2023, 177 providers representing all 29 MCM programs completed the survey (52% of 341 eligible providers and 88.5% of 200, the target sample size). About half (52%) of participants were 40–59 years old, the majority (72%) identified as women, and the plurality (41%) identified as Latino/a. Nearly three-quarters of participants (70%) had been providing services through a RWPA MCM program for >2 years, and the majority were either in a patient navigator-type role (38%) or in a case manager or care coordinator-type role (26%), which reflects staffing patterns at MCM/Care Coordination programs in NYC/Tri-County (data not shown). We chose the two-group LCA solution because we judged it to balance interpretability and model fit statistics (see Supplemental Table 3). The only statistically significant differences in participant characteristics by LCA group were gender identity and having heard about LAI ART before the survey from other MCM program staff. More Group 1 than Group 2 participants identified as women (82% vs 68%, p=0.018) and more Group 1 participants had heard of LAI ART before the survey from MCM program staff (68% vs. 47%, p=0.017). (See Table 1.)

**Table 1.**
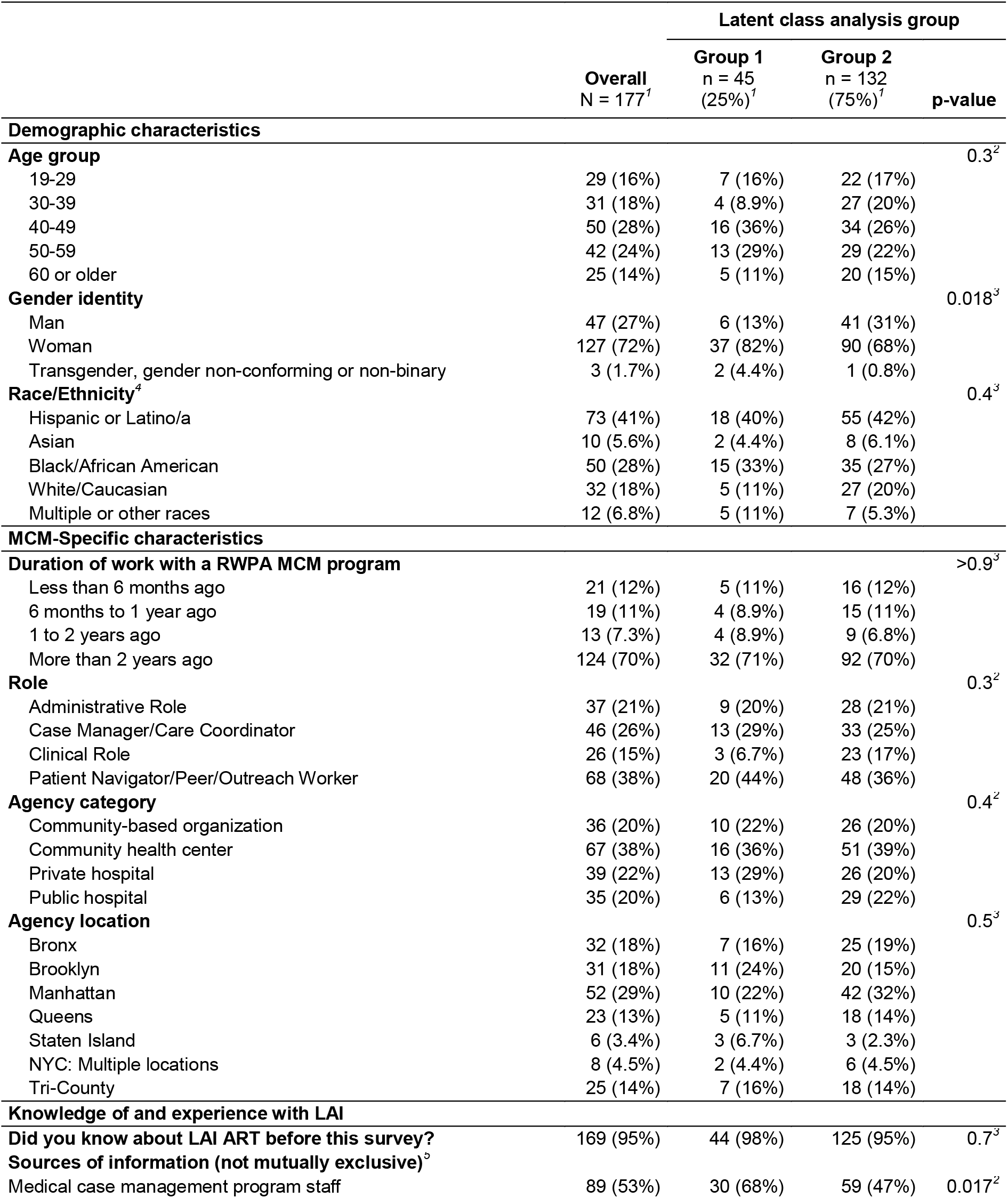
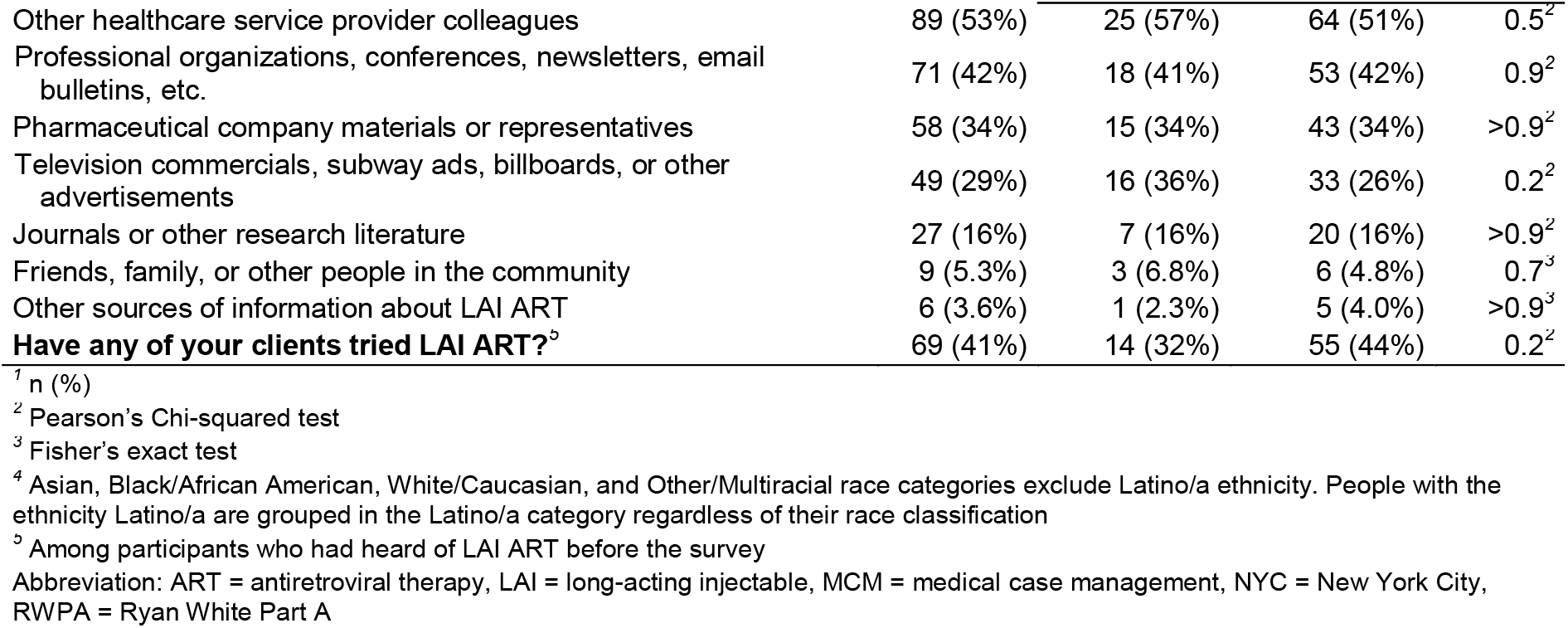
Demographics, medical case management-specific characteristics, and knowledge of long-acting injectable ART, APPLI Provider DCE.

### Measures of influence & preference

Overall and for Group 2 (n=132), Type of ART Medication was the most influential attribute (relative importance 32% [95% CI: 30%, 34%] and 38% [95% CI: 38%, 39%], respectively), followed by Rewards for Clients (31% [95% CI: 30%, 33%] and 37% [95% CI: 37%, 37%], respectively), Support for Clients (21% [95% CI: 19%, 22%] and 14% [95% CI: 14%, 15%], respectively), and Service Location and Mode (16% [95% CI: 15%, 17%] and 11% [95% CI: 11%, 11%], respectively). However, for Group 1 (n=45), Support for Clients (40% [95% CI: 38%, 41%]) and Service Location and Mode (32% [95% CI: 30%, 33%]) were the most important attributes. (See Supplemental Table 4 and Figure 1.)

**Figure 1.**
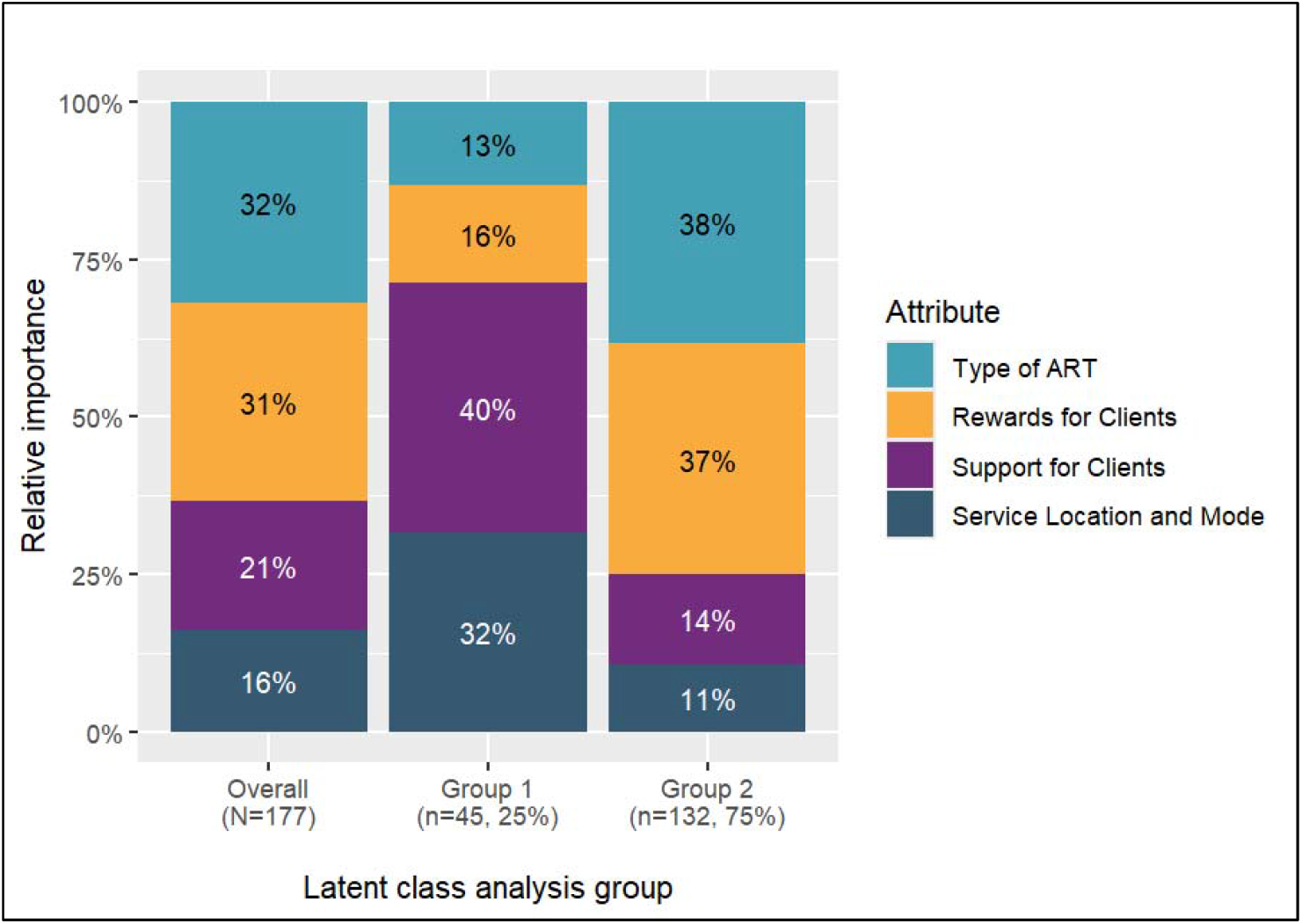
Mean relative importances of attributes^*1*^ by latent class analysis group, APPLI Provider DCE Abbreviations: APPLI = Assessing Perceptions and Preferences Around Long-acting Injectables; ART = antiretroviral therapy DCE = discrete choice experiment. ^*1*^ Attributes are sorted in descending order of relative importance within Group 2, the larger of the two latent class analysis groups.

“Bimonthly LAI ART” was the preferred Type of ART Medication for Group 1 (utility 26 [95% CI: 23, 30]). In Service Location and Mode, Group 1 participants preferred programs with “Injections at clinic by appointment” (66 [95% CI: 63, 69]); in Support for Clients, they preferred “Calls and texts to remind about injection appointments” (67 [95% CI: 64, 69]) or “Post-injection follow-up calls” (47 [95% CI: 44, 51]); and in Rewards for Clients, they preferred “$20 gift card for on-time injection visits” (37 [95% CI: 35, 39]). They had a low preference for the None option (-79 [95% CI: -106, -53]).

Group 2 participants also preferred “Bimonthly LAI ART” for Type of ART Medication, but they had a stronger preference (77 [95% CI: 76, 77]) than Group 1 participants. Group 2 participants preferred “Injections at home by appointment” for Service Location and Mode (18 [95% CI: 17, 19]); and “Free transportation to get injections” in Support for Clients (30 [95% CI: 30, 31]). Similar to Group 1, Group 2 preferred “$20 gift card for on-time injection visits” (63 [95% CI: 63, 64]) in Rewards for Clients, but they had a stronger preference for this level than Group 1. Group 2 participants had a very low preference for the None option (-495 [95% CI: - 499, -491]). (See Supplemental Table 4 and Figure 2.)

**Figure 2.**
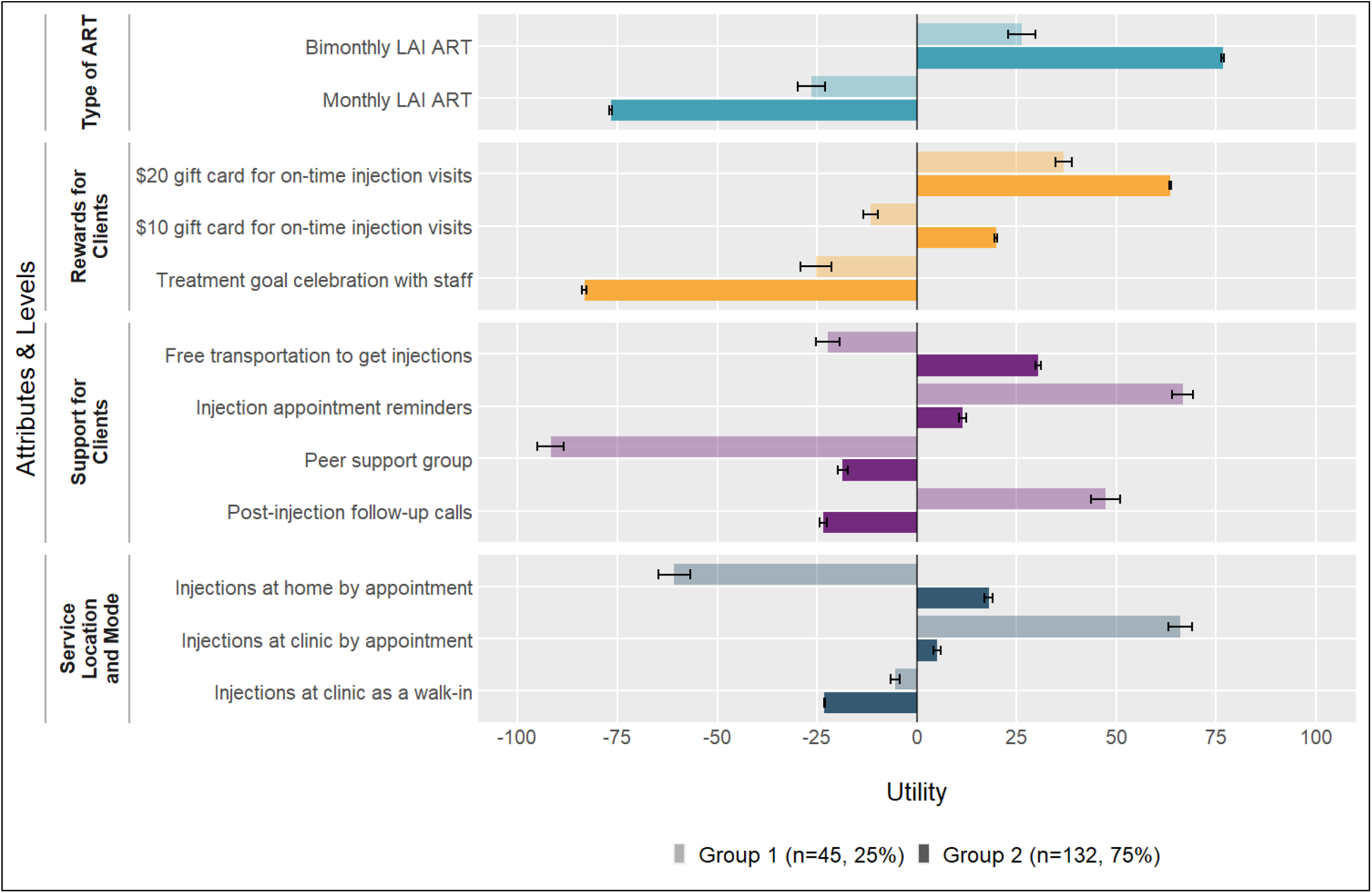
Mean preferences^*1*^ (with 95% CI) for type of ART and related supports by latent class analysis group, APPLI Provider DCE Abbreviations: APPLI = Assessing Perceptions and Preferences Around Long-acting Injectables; CI = confidence interval; DCE = discrete choice experiment; LAI ART = long-acting injectable antiretroviral therapy. ^*1*^ Attributes and levels sorted within attribute in descending order of preference (utility) within Group 2, the larger of the two latent class analysis groups.

### Ranking & motivations

The levels were ranked in the same order by utility from the DCE and in the independent ranking exercise, overall, with some differences between the ranking methods within the LCA groups (See Supplemental Table 5).

Participants then ranked their motivations for the order in which they independently ranked the levels. Overall, “Maximizing adherence and viral suppression” was in 76% of participants’ top-3 rankings, “Matching clients’ preferences” was in 69% of participants’ top-3 rankings, “Making it easy for clients to manage” was in 66% of participants’ top-3 rankings, and “Minimizing missed appointments” was in 51% of participants’ top-3 rankings. These four motivations far outranked the options related to administrative or financial feasibility for the agency and staff job satisfaction (see Supplemental Table 6). Rankings were similar across latent class groups and roles (data not shown).

### Implementation measures

The three implementation measures were each averaged across four items from 1 (“Completely disagree”) to 5 (“Completely agree”) to create a score for each participant. There was evidence of skew in the AIM, with medians less than means. The overall median scores were 4 (IQR 4, 5) on the AIM, 4 (IQR 3.75, 5) on the IAM, and 4 (IQR 3.75, 4.5) on the FIM, and the group-specific medians were similar (see Table 2). However, the overall mean scores were 4.2 (95% CI 4.1, 4.3) on the AIM, 4.02 (95% CI 3.9, 4.1) on the IAM, and 3.99 (95% CI 3.9, 4.1) on the FIM, with slightly higher scores on the AIM in Group 2 (4.22 [95% CI 4.1, 4.3]) compared to Group 1 (4.14 [95% CI 3.9, 4.4]). Though the median and mean scores were not statistically significantly different across latent class groups, both the median and mean AIM scores were statistically significantly higher than the IAM and the FIM scores overall and in Group 2. Supplemental Figure 2 plots the percent of participants who endorsed “Agree” or Completely Agree” for each item, showing consistently higher values in Group 2 than Group 1.

**Table 2.** Implementation measure scores^*1*^ overall and by latent class analysis group.

| Implementation measures | Overall<br>N = 177 | Latent class analysis groups |  | p-value<sup>2</sup> |
| --- | --- | --- | --- | --- |
|  |  | Group 1<br>n = 45 (25%) | Group 2<br>n = 132 (75%) |  |
| Acceptability of Intervention Measure (AIM) |  |  |  | 0.6 |
| Mean (95% CI) | 4.20 (4.1, 4.3) | 4.14 (3.9, 4.4) | 4.22 (4.1, 4.3) |  |
| Median (Q1, Q3) | 4.00 (4.00, 5.00) | 4.00 (4.00, 4.75) | 4.00 (4.00, 5.00) |  |
| Intervention Appropriateness Measure (IAM) |  |  |  | >0.9 |
| Mean (95% CI) | 4.02 (3.9, 4.1) | 4.02 (3.8, 4.3) | 4.02 (3.9, 4.2) |  |
| Median (Q1, Q3) | 4.00 (3.75, 5.00) | 4.00 (3.25, 5.00) | 4.00 (3.75, 4.63) |  |
| Feasibility of Intervention Measure (FIM) |  |  |  | >0.9 |
| Mean (95% CI) | 3.99 (3.9, 4.1) | 4.03 (3.8, 4.2) | 3.98 (3.9, 4.1) |  |
| Median (Q1, Q3) | 4.00 (3.75, 4.50) | 4.00 (3.50, 4.50) | 4.00 (3.75, 4.25) |  |

<sup>1</sup> Each measure is the average score of four related questions scored from 1 (“Completely disagree”) to 5 (“Completely agree”)
<sup>2</sup> Wilcoxon rank sum test

### Sensitivity analyses excluding poor quality responses

In the first sensitivity analysis (n=168), we excluded one participant who exhibited straight-lining behavior and eight participants whose DCE completion times were within the fastest 5^th^ percentile. In the second (n=160), we also excluded the one participant who exhibited straight-lining, and 16 participants whose completion times were within the fastest 10^th^ percentile. In the first sensitivity analysis, the differences in relative importances and utilities ranged from -0.2 percentage points (pp) to 2.4pp and from -8 utiles to 6 utiles, respectively. In the second sensitivity analysis, the differences in relative importances and utilities ranged from -6.4pp to 4.5pp and from -16 utiles to 14 utiles, respectively. In spite of these quantitative changes, the main findings from the original analysis remained unchanged. See Supplemental Materials and Supplemental Tables 7 and 8.

## Discussion

We investigated RWPA MCM provider preferences for MCM services and supports for clients interested in LAI ART. In the two-group LCA solution, Group 1 had a higher proportion of women than Group 2, and Group 1 members reported knowledge of LAI ART from MCM program staff more frequently than Group 2. Group 1 had higher preferences for more traditional service locations, modes, and supports such as injections at a clinic by appointment, injection appointment reminders and post-injection follow-up calls—similar to what is and has been done to support adherence to daily pill ART—whereas Group 2 had higher preferences for more bespoke and client-centered support for adherence, in the form of transportation to injection appointments and the option for clients to receive injections at home. Though the strength of the preferences varied between groups, both groups preferred bimonthly over monthly injections, and had low preferences for peer support groups and treatment goal celebrations with staff, given Reward options that provided more material utility to clients.

These results align with existing findings both from the client/patient perspective as well as the provider perspective. Results from the APPLI client DCE show that participants tended to prefer less frequent injections, delivery of pills or injections at home, transportation to appointments when needed, and to have similar preferences for gift cards over celebrations with staff (manuscript under review). Preferences for less frequent injections and receiving services at home has also been found in the emerging LAI preference literature among both patients/clients and providers,^33,34,51-53^ suggesting that client and provider preferences may be concordant for features that enhance the convenience of and access to this new treatment modality. This is supported by the finding that participants ranked motivations centering on the client experience higher than motivations pertaining to administrative or financial feasibility. Both LCA groups had negative utilities for the None option, which indicates that they found at least one of the LAI ART options preferrable to none.

Participants agreed in both groups that LAI ART was acceptable, appropriate, and feasible, with higher endorsement of acceptability, on average, than the other implementation measures. The implementation measure results illustrate a pattern consistent with Group 2 being generally more open than Group 1 to make changes in their practices that might be needed to support access and adherence to LAI ART. Previous research has also found high ratings of the appropriateness and acceptability of LAI ART, but lower feasibility.^31-33^ This survey did not directly assess readiness in terms of agency staffing, information technology, medication storage, and other resource allocation—concerns identified in previous research,^31-33^ but which would not be applicable to all agencies in the current study, as some do not directly provide clinical services. However, the high degree of openness to integrating supports for LAI ART indicated in our results bodes well for the actual provision of supports for LAI ART through MCM programs.

Our study has the following limitations. We may have omitted client-, provider-, or agency-level characteristics that would have enabled us to distinguish the groups of providers (or their clients and agencies) in order to provide context to the observed preference heterogeneity and understand better how to tailor the implementation of client supports for LAI ART in different contexts. We may not have included attributes and levels that captured the service features that were the most important or preferable to providers, in spite of the inclusion of providers in formative DCE design work through focus groups and Advisory Board meetings. Lacking routine data collection specific to treatment supports for clients on LAI ART, we were unable to determine what agencies are currently doing to facilitate LAI ART access and adherence among clients electing to initiate it.

## Conclusion

The engagement and buy-in of MCM providers in the integration of LAI ART into existing ART support programs may reduce observed disparities in VS by ensuring meaningful client access to and informed decision-making regarding their treatment options. Our implementation science-focused study suggests that providers of MCM services in NYC and the Tri-County area endorse offering a range of services to support clients’ access to this newer treatment option, and are motivated by factors related to clients’ convenience and adherence.

## Data availability statement

De-identified discrete choice experiment data only will be made available upon reasonable request.

## Ethics approval statement

The study methods were reviewed and approved by the Institutional Review Board of the New York City Department of Health and Mental Hygiene, protocol 20-096.

## Patient consent statement

Clients were required to review a consent form and provide electronic consent before they could begin the survey.

## Permission to reproduce material from other sources

N/A

## Clinical trial registration

NCT05833542

## Acknowledgements

We gratefully acknowledge the contributions of Aimee Campbell, Chunki Fong, Gina Gambone, Honoria Guarino, Grace Herndon, Javier Lopez-Rios, Monique Millington, Tyeirra Seabrook, the Assessing Perceptions and Preferences Around Long-acting Injectables (APPLI) study advisory board, and all of our Ryan White Part A medical case management (MCM) program sites and their clients. The study benefitted from the support of the HIV Health and Human Services Planning Council of New York, the New York City Health Department HIV Care and Treatment Program leadership and quality management staff and the Einstein-Rockefeller-CUNY Center for AIDS Research.

