## Supplemental materials for "Exploring provider preferences in the design of HIV treatment packages integrating long-acting injectable antiretroviral therapy in New York Ryan White Part A medical case management programs: A discrete choice experiment"

**Supplemental Table 1. Attributes and levels with helper images**

| Attribute | Level | Level abbreviation for figures and text references | Helper image |
|---|---|---|---|
| Type of ART Medication | An ART injection in each buttock **once a month** | Monthly LAI ART | 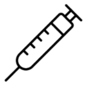 |
| | An ART injection in each buttock **once every two months** | Bimonthly LAI ART | 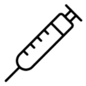 |
| Service Location and Mode | Injections **by appointment** at clinic | Injections by appointment at clinic | 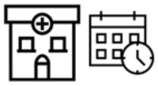 |
| | Walk-in for injections at clinic with **no appointment required** | Injections as a walk-in at clinic | 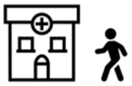 |
| | Injections **by appointment** at home* | Injections by appointment at home | 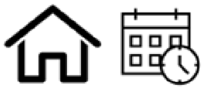 |
| Support for Clients | Calls and texts to remind about injection appointments | Injection appointment reminders | 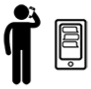 |
| | Free transportation provided to get injections* | Free transportation to get injections | 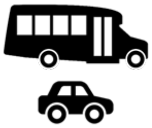 |
| | Post-injection follow-up calls from program staff | Post-injection follow-up calls | 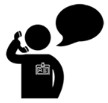 |
| | Peer support group for clients on or considering long-acting injectable ART | Peer support group | 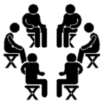 |
| Rewards for Clients | $10 gift card for every injection visit in the right time frame | $10 gift card for on-time injection visits | 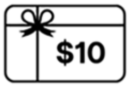  |
| | $20 gift card for every injection visit in the right time frame | $20 gift card for on-time injection visits | 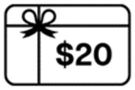  |
| | Rewards ceremony with program staff to celebrate meeting treatment goals | Treatment goal celebration with staff | 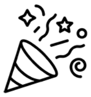 |

*Injections by appointment at home in the Service Location and Mode attribute was prohibited from appearing in an alternative with Free transportation provided to get injections in the Support for Clients attribute.
Abbreviations: ART=antiretroviral therapy

**Supplemental Figure 1. Sample task**

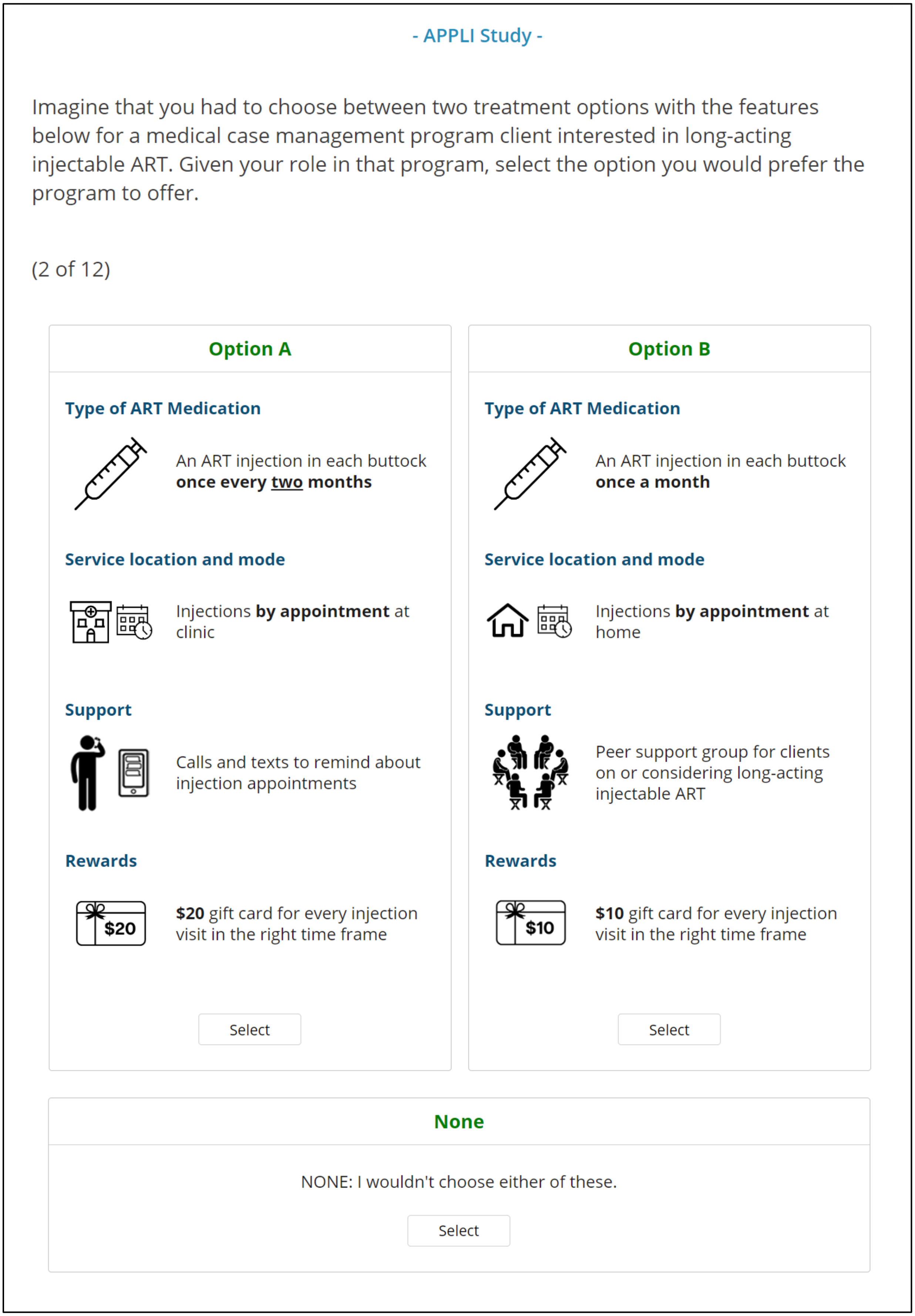

**Supplemental Table 2. Checklist for reporting discrete choice experiments in health**[1]

| Section Item | | Section and paragraph[2] |
|---|---|---|
| Purpose and rationale | | |
| 1 | Describe the real-world context and decision-maker that the hypothetical choice context seeks to replicate or inform | Introduction paragraphs 3 and 5 |
| 2 | Provide a rationale for using a DCE to answer the research question | Introduction paragraph 5 |
| Attributes and levels | | |
| 3 | Describe how attributes and levels were derived (e.g. literature review, interviews, focus groups, expert input) | Methods paragraph 1 |
| 4 | Provide the final list of attributes and levels | Supplemental Table 1 |
| Experimental design | | |
| 5 | Report the number of alternatives per choice set and whether they were labelled or unlabelled | Methods, Survey design and pilot testing, paragraph 3 |
| 6 | Describe response options (e.g. forced choice, opt-out, status quo) | Methods, Survey design and pilot testing, paragraph 3 |
| 7 | Describe the type of experimental design (e.g. orthogonal, D-efficient, Bayesian efficient, partial profile) | Methods, Survey design and pilot testing, paragraph 3 |
| 8 | Describe which effects are identified in the design (e.g. main effects, higher order interactions, functional form) | Methods, Statistical analysis, paragraph 1 |
| 9 | Describe the number of choice sets, blocks and choice sets per block | Methods, Survey design and pilot testing, paragraph 3 |
| 10 | Indicate how the experimental design was obtained (software, catalogue, other) | Methods, Survey design and pilot testing, paragraph 3 |
| Survey design | | |
| 11 | Provide a sample choice set and the instructions and background information given to respondents (e.g. providing the survey as an appendix) | Supplemental Figure 1 |
| 12 | Report any randomisation (e.g. choice set order, attribute order, alternative order, framing effects) | Methods, Survey design and pilot testing, paragraph 3 |
| 13 | Describe what was checked in piloting (e.g. understanding, respondent burden, timing, wording) | Methods, Survey design and pilot testing, paragraph 4 |
| 14 | Report whether information from the pilot was used to update the experimental design (e.g. priors, functional form of attributes) or survey design | Methods, Survey design and pilot testing, paragraph 4 |
| Sample and data collection | | |
| 15 | Report respondent inclusion/exclusion criteria | Methods, Study population and sample size, paragraph 1 |
| 16 | Describe how data were collected (e.g. mail, personal interview, web survey) | Methods, Recruitment & data collection, paragraph 1 |
| 17 | Report the response rate or cooperation rate, if possible | Results, Participant characteristics, paragraph 1 |
| 18 | Report the final sample size and how the sample size was determined | Methods, Study population and sample size, paragraph 2 |
| 19 | Describe respondent characteristics and representativeness of target population, if known | Results, Participant characteristics, paragraph 1 and Table 1 |
| Econometric analysis | | |
| 20 | Indicate coding of data (e.g. effects, dummy, continuous) including definitions | Methods, Statistical analysis, paragraph 1 |
| 21 | Report whether any respondents were removed and why (e.g. suspected fraudulent responses, rationality tests) | Methods, Statistical analysis, paragraph 1<br>Results, Sensitivity analyses excluding poor quality responses, paragraph 1<br>Supplemental materials |

| Section Item | | Section and paragraph[2] |
|---|---|---|
| 22 | Provide the rationale for model choice (e.g. conditional logit, mixed logit, latent class) and assumptions (e.g. error variance) | Methods, Statistical analysis, paragraph 1 |
| 23 | Report model specification | Methods, Statistical analysis, paragraph 1 |
| Reporting of results | | |
| 24 | Report the model performance, goodness of fit (if comparing models) | Supplemental Table 3 |
| 25 | Describe methods used for analysis of model results (e.g. calculation of marginal rate of substitution, attribute relative importance, welfare gain) | Methods, Statistical analysis, paragraph 1 |
| 26 | Report measures of precision for the output(s) of interest (e.g. confidence intervals) and how these were derived | Methods, Statistical analysis, paragraph 1 |

[1] From Ride et al., "A Reporting Checklist for Discrete Choice Experiments in Health: The DIRECT Checklist." PharmacoEconomics 2024

[2] Revised from "Page and paragraph" to "Section and paragraph" due to expected formatting changes between submitted in Microsoft Word and final formatting of published manuscript.

**Supplemental Table 3. Goodness of fit statistics of latent class analysis solutions**

| Groups | Number of participants | | Log-likelihood | Difference in log-likelihood | Percent Certainty† | Difference in percent certainty | AIC | Difference in AIC |
|---|---|---|---|---|---|---|---|---|
| | In smallest group | In largest group | | | | | | |
| 1 | 177 | - | -1839.22 | - | 21.18% | - | 3696.45 | - |
| 2 | 45 | 132 | -1718.25 | 120.97 | 26.36% | 5.18% | 3474.5 | -221.95 |
| 3 | 38 | 100 | -1658.27 | 59.98 | 28.93% | 2.57% | 3374.54 | -99.96 |
| 4 | 39 | 52 | -1608.05 | 50.22 | 31.09% | 2.16% | 3294.1 | -80.44 |
| 5 | 12 | 47 | -1566.77 | 41.28 | 32.86% | 1.77% | 3231.54 | -62.56 |

† Percent certainty between 0.2 and 0.4 indicates a good model fit (Hauber et al. 2016).

**Supplemental Table 4. Relative importances and utilities overall and by latent class analysis group**

| | | Overall<br>N = 177 | Latent class analysis groups[1]<br>Group 1<br>n = 45 (25%) | Group 2<br>n = 132 (75%) |
|---|---|---|---|---|
| **Attributes[2] (Mean relative importances [95% CI])** | | | | |
| Type of ART Medication | | 32% (30%, 34%) | 13% (11%, 15%) | 38% (38%, 39%) |
| Rewards for Clients | | 31% (30%, 33%) | 16% (14%, 17%) | 37% (37%, 37%) |
| Support for Clients | | 21% (19%, 22%) | 40% (38%, 41%) | 14% (14%, 15%) |
| Service Location and Mode | | 16% (15%, 17%) | 32% (30%, 33%) | 11% (11%, 11%) |
| **Levels[2] (Mean utilities [95% CI])** | | | | |
| Type of ART Medication | An ART injection in each buttock once every two months | 64 (61, 67) | 26 (23, 30) | 77 (76, 77) |
| | An ART injection in each buttock once a month | -64 (-67, -61) | -26 (-30, -23) | -77 (-77, -76) |
| Rewards for Clients | $20 gift card for every injection visit in the right time frame | 57 (55, 58) | 37 (35, 39) | 63 (63, 64) |
| | $10 gift card for every injection visit in the right time frame | 12 (9.8, 14) | -12 (-13, -9.7) | 20 (19, 20) |
| | Rewards ceremony with program staff to celebrate meeting treatment goals | -69 (-72, -65) | -25 (-29, -21) | -83 (-84, -83) |
| Support for Clients | Free transportation provided to get injections | 17 (14, 21) | -22 (-25, -19) | 30 (30, 31) |
| | Calls and texts to remind about injection appointments | 25 (22, 29) | 67 (64, 69) | 11 (11, 12) |
| | Peer support group for clients on or considering LAI ART | -37 (-42, -32) | -92 (-95, -88) | -19 (-20, -17) |
| | Post-injection follow-up calls from program staff | -5 (-10, -0.68) | 47 (44, 51) | -23 (-24, -22) |
| Service Location and Mode | Injections at home by appointment | -2 (-7.3, 3.3) | -61 (-65, -57) | 18 (17, 19) |
| | Injections at clinic by appointment | 21 (16, 25) | 66 (63, 69) | 5 (4.1, 6.0) |
| | Walk-in for injections at clinic with no appointment required | -19 (-20, -17) | -5 (-6.5, -4.2) | -23 (-23, -23) |
| NONE: I wouldn't choose either of these[3] | | -389 (-417, -361) | -79 (-106, -53) | -495 (-499, -491) |

Abbreviation: ART = antiretroviral therapy; CI = confidence interval; LAI ART = long-acting injectable ART
[1] All comparisons across latent class groups are statistically significant at $p<0.001$ by the Wilcoxon rank sum test
[2] Attributes and levels are sorted in descending order of relative importance or utility, respectively within Group 2, the larger of the two latent class analysis groups.
[3] The utility for the NONE option is an alternative-specific constant.

**Supplemental Table 5. Independent ranking of levels for all participants (N=177)**

| | Overall (N=177) | | | | Latent class analysis group: Group 1 (n=45) | | | | Group 2 (n=132) | | | |
|---|---|---|---|---|---|---|---|---|---|---|---|---|
| **Attributes and Levels** | 1 | 2 | 3 | 4 | 1 | 2 | 3 | 4 | 1 | 2 | 3 | 4 |
| **Type of ART** | | | | | | | | | | | | |
| Bimonthly LAI ART | **151** | 26 | | | **34** | 11 | | | **117** | 15 | | |
| Monthly LAI ART | 26 | **151** | | | 11 | **34** | | | 15 | **117** | | |
| **Service Location and Mode** | | | | | | | | | | | | |
| Injections at clinic by appointment | **72** | 55 | 50 | | **26** | 13 | 6 | | **46** | 42 | 44 | |
| Injections at clinic as a walk-in | 57 | **65** | 55 | | 11 | **15** | **19** | | **46** | **50** | 36 | |
| Injections at home by appointment | 48 | 57 | **72** | | 8 | **17** | **20** | | 40 | 40 | **52** | |
| **Support for Clients** | | | | | | | | | | | | |
| Injection appointment reminders via calls & texts | **103** | 50 | 16 | 8 | **30** | 9 | 5 | 1 | **73** | 41 | 11 | 7 |
| Free transportation to get injections | 55 | **74** | 29 | 19 | 11 | **17** | 10 | 7 | 44 | **57** | 19 | 12 |
| Post-injection follow-up calls | 10 | 35 | **100** | 32 | 3 | **16** | **23** | 3 | 7 | 19 | **77** | 29 |
| Peer support for LAI ART | 9 | 18 | 32 | **118** | 1 | 3 | 7 | **34** | 8 | 15 | 25 | **84** |
| **Rewards for Clients** | | | | | | | | | | | | |
| $20 gift card for on-time injection visits | **144** | 23 | 10 | | **32** | 11 | 2 | | **112** | 12 | 8 | |
| $10 gift card for on-time injection visits | 16 | **135** | 26 | | 6 | **31** | 8 | | 10 | **104** | 18 | |
| Treatment goal celebration with staff | 17 | 19 | **141** | | 7 | 3 | **35** | | 10 | 16 | **106** | |

**Supplemental Table 6. Ranking of motivations for independent level rankings for all participants (N=177)**

| | Rank | | | |
|---|---|---|---|---|
| **Motivation** | 1 | 2 | 3 | Total |
| Maximizing adherence and viral suppression | 55 (31%) | 42 (24%) | 38 (21%) | 135 (76%) |
| Matching clients' preferences | 60 (34%) | 38 (21%) | 24 (14%) | 122 (69%) |
| Making it easy for clients to manage | 35 (20%) | 40 (23%) | 42 (24%) | 117 (66%) |
| Minimizing missed appointments | 15 (8%) | 40 (23%) | 36 (20%) | 91 (51%) |
| Aligning with our program priorities/organizational mission | 7 (4%) | 6 (3%) | 13 (7%) | 26 (15%) |
| Making it easy for staff to implement | 3 (2%) | 4 (2%) | 9 (5%) | 16 (9%) |
| Doing what is administratively feasible in my program setting | 1 (1%) | 3 (2%) | 5 (3%) | 9 (5%) |
| Doing what is financially feasible in my program setting | 1 (1%) | 3 (2%) | 4 (2%) | 8 (5%) |
| Enhancing job satisfaction for staff | 0 (0%) | 1 (1%) | 5 (3%) | 6 (3%) |
| Other | 0 (0%) | 0 (0%) | 1 (1%) | 1 (1%) |
| Total | 177 | 177 | 177 | 531 |

**Supplemental Figure 2. Percent agree or strongly agree with individual implementation measure items**

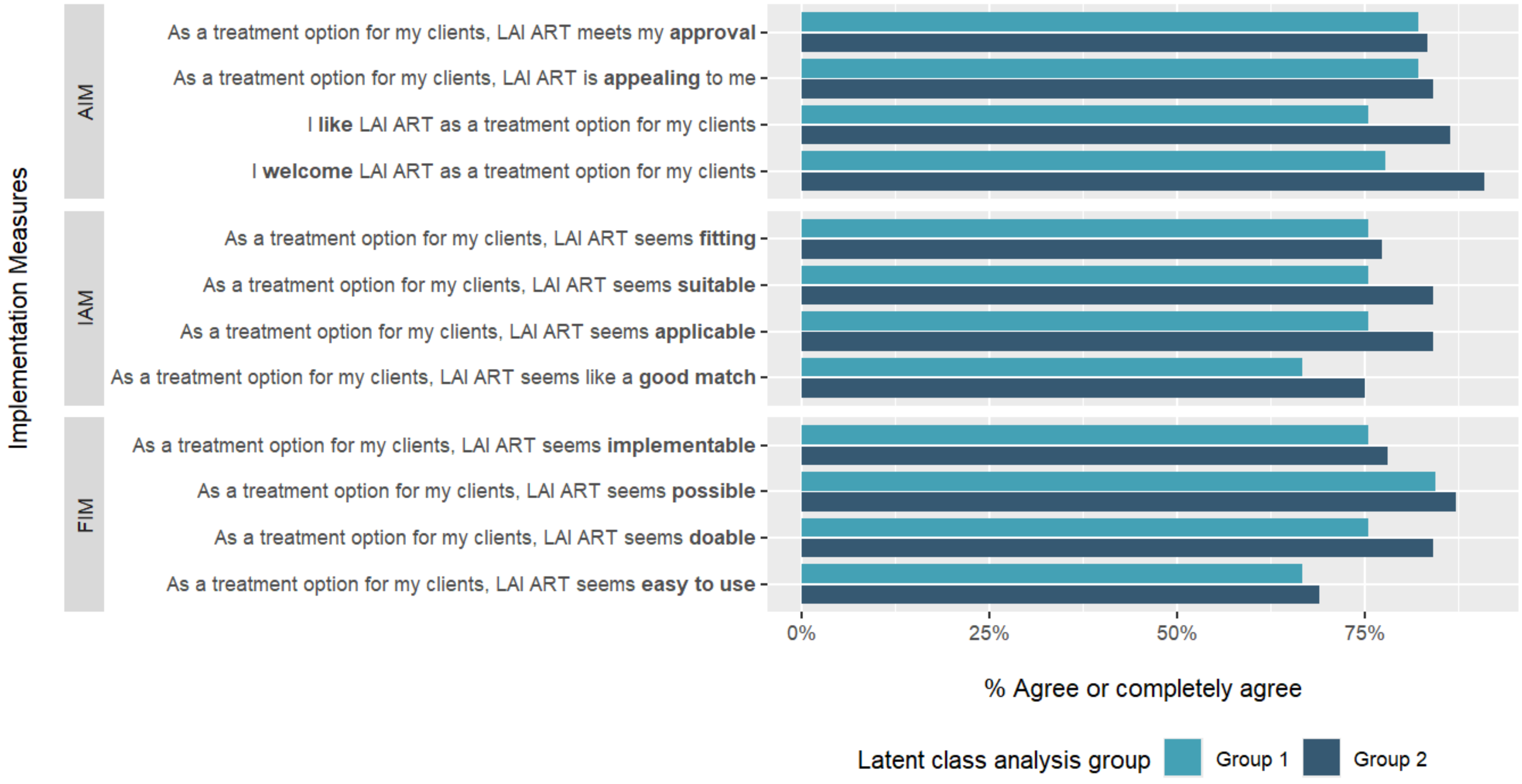


Abbreviations: AIM=acceptability of intervention measure, IAM= intervention appropriateness measure, FIM=feasibility of intervention measure, LAI ART=long-acting injectable antiretroviral therapy.

**Sensitivity analyses excluding poor quality responses**

In the first sensitivity analysis (n=168), two participants were dropped from Group 1, seven participants were dropped from Group 2, and three participants moved from Group 2 to Group 1, leaving 46 participants in Group 1 and 122 participants in Group 2. Overall, the values of relative importance were within one percentage point of the original analysis, and utilities were within two utiles of the original analysis. The ordering of the attributes, in terms of relative importance, and of the levels, in terms of utilities, remained unchanged. In Group 1, due to two percentage point changes in the relative importance values, Type of ART became the third most important attribute (instead of fourth) and Rewards for Clients became the fourth most important attribute (instead of third), switching places in the attribute ordering. All levels were ordered the same as in the original analysis. The ranges of utilities in Group 1 became slightly more extreme within all attributes, and the utility for the none/opt-out option became more negative. In Group 2, the attributes and levels were ordered the same as in the original analysis, and the differences in relative importances and utilities were small; the utility for the none/opt-out option became more negative, as in Group 1. See Supplemental Table 6.

In the second sensitivity analysis (n=160), four participants were dropped from Group 1, 13 participants were dropped from Group 2, and two participants moved from Group 1 to group 2, leaving 39 participants in Group 1 and 119 participants in Group 2. As in the first sensitivity analysis, overall, the ordering of the attributes and of the levels remained unchanged. In Group 1, as above, Type of ART and Rewards for Clients switched places in the attribute ordering. There were larger changes in the relative importances and utilities across all attributes in Group 1 than in the first sensitivity analysis, though all levels were ordered the same as in the original analysis. The utility for the none/opt-out option became less negative. In Group 2, Type of ART became the most important attribute (instead of second) and Rewards for Clients became the second most important attribute (instead of first), due to one percentage point changes in relative importance. There were small changes in utilities, and the utility for the none/opt-out option became more negative. See Supplemental Table 7.

**Supplemental Table 7. Sensitivity analysis relative importances and utilities overall and by latent class group, excluding responses exhibiting straight-lining and completion times within the fastest 5th percentile (N=168)**

| | | | Latent class analysis groups[1] | |
|---|---|---|---|---|
| | | **Overall** (N = 168) | **Group 1** (n = 46, 27%) | **Group 2** (n = 122, 73%) |
| **Attributes[2] (Mean relative importances [95% CI])** | | | | |
| Type of ART Medication | | 33% (31%, 34%) | 15% (14%, 17%) | 39% (39%, 39%) |
| Rewards for Clients | | 31% (29%, 32%) | 14% (13%, 16%) | 37% (37%, 37%) |
| Support for Clients | | 20% (18%, 22%) | 39% (38%, 41%) | 13% (13%, 13%) |
| Service Location and Mode | | 16% (15%, 18%) | 31% (29%, 33%) | 11% (11%, 11%) |
| **Levels[2] (Mean utilities [95% CI])** | | | | |
| Type of ART Medication | An ART injection in each buttock once every two months | 65 (62, 69) | 31 (28, 34) | 78 (78, 79) |
| | An ART injection in each buttock once a month | -65 (-69, -62) | -31 (-34, -28) | -78 (-79, -78) |
| Rewards for Clients | $20 gift card for every injection visit in the right time frame | 55 (53, 57) | 31 (29, 33) | 64 (64, 64) |
| | $10 gift card for every injection visit in the right time frame | 13 (11, 15) | -5 (-6.8, -4.0) | 20 (20, 20) |
| | Rewards ceremony with program staff to celebrate meeting treatment goals | -68 (-72, -64) | -26 (-30, -22) | -84 (-84, -84) |
| Support for Clients | Free transportation provided to get injections | 16 (13, 19) | -16 (-18, -13) | 28 (28, 28) |
| | Calls and texts to remind about injection appointments | 26 (22, 30) | 65 (62, 67) | 11 (11, 12) |
| | Peer support group for clients on or considering LAI ART | -39 (-44, -34) | -92 (-96, -89) | -19 (-20, -18) |
| | Post-injection follow-up calls from program staff | -3 (-7.5, 1.4) | 44 (40, 47) | -21 (-21, -20) |
| Service Location and Mode | Injections at home by appointment | -2 (-7.0, 3.6) | -57 (-61, -54) | 19 (18, 20) |
| | Injections at clinic by appointment | 21 (17, 26) | 67 (64, 70) | 4 (3.0, 4.7) |
| | Walk-in for injections at clinic with no appointment required | -20 (-20, -19) | -10 (-11, -9.0) | -23 (-23, -23) |
| NONE: I wouldn't choose either of these[3] | | -391 (-420, -362) | -91 (-115, -66) | -504 (-506, -502) |

Abbreviation: ART = antiretroviral therapy; CI = confidence interval; LAI ART = long-acting injectable ART

[1] All comparisons across latent class groups are statistically significant at p<0.001 by the Wilcoxon rank sum test

[2] Attributes and levels are sorted in descending order of relative importance or utility, respectively within Group 2, the larger of the two latent class analysis groups.

[3] The utility for the NONE option is an alternative-specific constant.

**Supplemental Table 8. Sensitivity analysis relative importances and utilities overall and by latent class group, excluding responses exhibiting straight-lining and completion times within the fastest 10th percentile (N=160)**

| | | Overall (N = 160) | Latent class analysis groups[1] Group 1 (n = 39, 24%) | Group 2 (n = 121, 76%) |
|---|---|---|---|---|
| **Attributes[2] (Mean relative importances [95% CI])** | | | | |
| Rewards for Clients | | 32% (30%, 34%) | 11% (9.5%, 12%) | 39% (38%, 39%) |
| Type of ART | | 33% (32%, 35%) | 19% (18%, 20%) | 38% (38%, 38%) |
| Support for Clients | | 18% (17%, 20%) | 36% (35%, 37%) | 13% (13%, 13%) |
| Service Location and Mode | | 16% (15%, 18%) | 35% (33%, 36%) | 10% (10%, 10%) |
| **Levels[2] (Mean utilities [95% CI])** | | | | |
| Rewards for Clients | $20 gift card for every injection visit in the right time frame | 57 (54, 60) | 26 (24, 29) | 67 (66, 67) |
| | $10 gift card for every injection visit in the right time frame | 14 (12, 16) | -9 (-11, -7.7) | 21 (21, 21) |
| | Rewards ceremony with program staff to celebrate meeting treatment goals | -71 (-76, -66) | -17 (-21, -13) | -88 (-89, -87) |
| Type of ART Medication | An ART injection in each buttock once every two months | 67 (64, 70) | 37 (35, 40) | 76 (76, 77) |
| | An ART injection in each buttock once a month | -67 (-70, -64) | -37 (-40, -35) | -76 (-77, -76) |
| Support for Clients | Free transportation provided to get injections | 13 (9.8, 17) | -27 (-30, -25) | 26 (26, 27) |
| | Calls and texts to remind about injection appointments | 26 (23, 29) | 62 (60, 64) | 14 (13, 15) |
| | Post-injection follow-up calls from program staff | -3 (-7.8, 1.3) | 47 (44, 50) | -19 (-20, -18) |
| | Peer support group for clients on or considering LAI ART | -36 (-40, -32) | -81 (-84, -79) | -22 (-23, -20) |
| Service Location and Mode | Injections at home by appointment | -7 (-13, -0.76) | -71 (-75, -68) | 14 (13, 16) |
| | Injections at clinic by appointment | 23 (19, 27) | 67 (64, 69) | 9 (8.4, 10) |
| | Walk-in for injections at clinic with no appointment required | -17 (-19, -15) | 5 (3.3, 6.2) | -24 (-24, -23) |
| NONE: I wouldn't choose either of these[3] | | -394 (-426, -363) | -49 (-73, -26) | -506 (-510, -501) |

Abbreviation: ART = antiretroviral therapy; CI = confidence interval; LAI ART = long-acting injectable ART

[1] All comparisons across latent class groups are statistically significant at $p<0.001$ by the Wilcoxon rank sum test

[2] Attributes and levels are sorted in descending order of relative importance or utility, respectively within Group 2, the larger of the two latent class analysis groups.

[3] The utility for the NONE option is an alternative-specific constant.
